# Brain mineralization in postoperative delirium and cognitive decline

**DOI:** 10.1101/2023.12.02.23299086

**Authors:** Florian Lammers-Lietz, Friedrich Borchers, Insa Feinkohl, Stefan Hetzer, Cicek Kanar, Gunnar Lachmann, Claudia Chien, Claudia Spies, Georg Winterer, Laszlo Zaborszky, Norman Zacharias, Friedemann Paul

## Abstract

Delirium is a severe postoperative complication associated with poor overall and especially neurocognitive prognosis. Altered brain mineralization is found in neurodegenerative disorders but has not been studied in postoperative delirium and postoperative cognitive decline. We hypothesized that mineralization-related hypointensity in susceptibility-weighted magnetic resonance imaging (SWI) is associated with postoperative delirium and cognitive decline.

We analyzed a subsample of cognitively healthy patients ≥65 years presenting for elective major surgery who underwent SWI before (N=65) and three months after surgery (N=33) as part of a subproject in the BioCog study. We measured relative SWI intensities in basal ganglia, hippocampus, and posterior basal forebrain cholinergic system (pBFCS). A post-hoc analysis of two pBFCS subregions (Ch4, Ch4p) was conducted. Patients were screened for delirium until the seventh postoperative day. Cognitive testing was performed before and three months after surgery.

Preoperative relative SWI hypointensities in the basal ganglia and pBFCS were associated with increased risk for postoperative delirium after adjustment for surgery duration. After additional adjustment for age, sex, preoperative MMSE and region volume, only the association of pBFCS hypointensity and postoperative delirium remained significant. Adjusted for surgery duration, perioperative change in relative SWI intensities of the pBFCS was associated with cognitive decline three months after surgery. This association remained at a trend level after adjustments for age, sex, and region volume, but a significant independent association especially with pBFCS-subregion Ch4p was found in a post-hoc analysis.

Brain mineralization, particularly in the cerebral cholinergic system, could be a pathomechanism in postoperative delirium and cognitive decline.

## 1 Introduction

Delirium is a common, but severe complication after surgery and anesthesia marked by acute disturbances in attention, awareness, cognition, psychomotor behavior, and emotion. Especially older patients are at high risk for delirium (Androsova et al., 2015) and in particular postoperative delirium (POD) will gain further relevance as life expectancy is increasing in Western societies, and the need for surgical interventions is higher among the older (Fowler et al., 2019; Nielsen et al., 2021). This challenges healthcare systems since POD is associated with poor neurocognitive outcome, hospitalization, treatment costs, re-institutionalization and mortality (Abelha et al., 2013; Drews et al., 2015; Hshieh et al., 2017; Inouye et al., 2016; Koebrugge et al., 2009; Peter et al., 2016; Robinson et al., 2009; Rudolph et al., 2008).

Pre-existing neurodegenerative diseases are a risk factor for POD (Dasgupta & Dumbrell, 2006), and vice versa, as cognitive decline is a common long-term sequela of POD in cognitively healthy individuals (Robinson et al., 2009; Teipel et al., 2018), but potentially shared pathomechanisms are yet to be investigated.

Cerebral mineralization, such as iron deposition and calcifications, have been suggested as a common feature in aging (Burgetova et al., 2021) and several neurodegenerative and neuroinflammatory diseases, such as Alzheimer’s disease (AD) (Damulina et al., 2020), Parkinson’s disease (PD) (Biondetti et al., 2021) and multiple sclerosis (Chawla et al., 2018; Chawla et al., 2016; Hametner et al., 2013). E.g., iron has been found to be associated with tau deposits amyloid plaques in AD (Madsen et al., 2020; Spotorno et al., 2020; Wu et al., 2023). α-synuclein as the main constituent of abnormal aggregates in PD and Lewy Body dementia (LBD) has been shown to mediate potentially neurotoxic calcium influx as well as iron-dependent cell membrane oxidation (Angelova et al., 2020) and vice versa, iron accelerates the fibril formation of α-synuclein (Bharathi et al., 2007). For multiple sclerosis, it has been suggested that iron is deliberated from the damaged myelin sheath and subsequently accumulates in microglia (Hametner et al., 2013). Furthermore, both iron and calcium deposition have been observed in animal models of traumatic brain injury and status epilepticus (Aggarwal et al., 2018; Schweser et al., 2019).

However, only few data have been published with regard to metals in the pathogenesis of delirium: A study conducted in N=19 patients eventually reported altered plasma iron levels in patients with delirium tremens, but overall results were inconclusive (Hemmingsen & Kramp, 1980). A recent analysis of the UK biobank data suggested an association of homozygosity of hemochromatosis-related p.C282Y and cerebral magnetic resonance imaging (MRI) measures indicative of iron deposition as well as incident delirium (Atkins et al., 2021). A retrospective epidemiological study and one small randomized controlled trial suggested that brain-penetrating calcium channel blockers may reduce the risk for incident delirium (Colbourne & Harrison, 2022; Li et al., 2017). Hence, there are little to no studies investigating the role of iron deposits, tissue calcification and brain mineralization in POD.

Susceptibility weighted imaging (SWI) has previously been used to study increased iron deposition and calcification in aging (Harder et al., 2008) and conditions such as Parkinson’s disease (Meijer et al., 2015; Schneider et al., 2016). In a previous work, we studied cerebral microbleeds in POD using SWI (Lachmann et al., 2019). Although the work did not find evidence for an association of microhemorrhages with a predisposition for POD, it is unknown if iron deposits or calcification from other sources with distinct distribution patterns, e.g., metal-binding protein aggregates or accumulation in microglia, could contribute to POD development. Here, we use the same cohort to study iron deposition and brain calcification in three regions of interest (ROI): the basal ganglia, the hippocampus, and the posterior basal forebrain cholinergic system (pBFCS).

Physiological (Haacke et al., 2005), age-related (Acosta-Cabronero et al., 2016; Betts et al., 2016; Burgetova et al., 2021) and neurodegenerative iron deposition is most prominent and well documented in the basal ganglia and subcortical grey matter (Damulina et al., 2020; Fu et al., 2021; Meijer et al., 2015; Schneider et al., 2016). Less consistently, hippocampal iron deposition has been reported in aging (Acosta-Cabronero et al., 2016) and AD (Kim et al., 2017; Small et al., 2000; Zhu et al., 2009). Due to previous research suggesting cholinergic deficiency in delirium (Androsova et al., 2015; Hshieh et al., 2008; Muller et al., 2019), we decided to include the pBFCS as a ROI as well.

We hypothesize that cerebral iron deposition and calcification, measured by relative SWI hypointensities in these three brain regions will associate with POD and cognitive decline after surgery. We expect that preoperative hypointensity will indicate patients with a predisposition for POD. After surgery and anesthesia, further hypointensity in these regions will be tested for associations with poor cognitive outcome three months after surgery.

## 2 Materials & Methods

### 2.1 Study design

We analysed data from a small subproject of the BioCog (Biomarker Development for Postoperative Cognitive Impairment in the Elderly, www.biocog.eu) study. The BioCog project is a cohort study aiming at identification of risk factors for POD and postoperative cognitive dysfunction (POCD) which has been conducted at the Charité-Universitätsmedizin Berlin, Germany, and the University Medical Centre Utrecht, Netherlands.

Patients aged ≥65 years presenting for elective surgery with an expected duration of ≥60 minutes were screened at the Charité anaesthesiologic outpatient department and included after obtaining written informed consent. Patients were excluded in case of positive screening for pre-existing major neurocognitive disorder defined as a Mini-Mental Status Examination (MMSE) score of ≤23 points (Anthony et al., 1982), any condition interfering with the neurocognitive assessment (severe hearing or visual impairment, neurological or psychiatric disease), unavailability for follow-up assessment (e.g., due to homelessness), participation in another interventional study during hospital stay, accommodation in an institution due to official or judicial order, as well as missing informed consent.

Before surgery, patients underwent an extensive clinical interview and comprehensive cognitive assessment and blood sampling. Eligible patients were additionally scheduled for magnetic resonance imaging (MRI). After surgery, patients were prospectively screened for postoperative delirium for up to seven days until discharge. Three months after surgery, patients were invited for a follow-up assessment including cognitive testing, MRI and blood sampling. As part of the subproject, an additional SWI sequence was added to the MRI protocol to study the association of microbleeds with POD and POCD. Methods and procedures have already been described extensively in previous publications (Heinrich et al., 2020; Lachmann et al., 2019; Lammers-Lietz et al., 2022; Lammers et al., 2018; Winterer et al., 2018).

All study procedures were conducted in line with the declaration of Helsinki with approval by the local medical ethics committees of the study centres in Berlin, Germany (EA2/092/14) and Utrecht, Netherlands (14-469). All participants gave written informed consent prior to inclusion in the study. The study is registered at clinicaltrials.gov under NCT02265263.

### 2.2 Screening for postoperative delirium

Delirium assessment was conducted independently from the routine hospital procedures by the study team twice daily starting on the day of surgery. POD was diagnosed in accordance with criteria in the 5th edition of Diagnostic and Statistical Manual of Mental Disorders (DSM-5) or a positive screening result on the Nursing Delirium Screening Scale (Nu-DESC, ≥ 2 points), either Confusion Assessment Method (CAM) score or CAM for the Intensive Care Unit (CAM-ICU) or written evidence for delirium found by patient chart review.

### 2.3 Assessment of cognitive function

Originally, POCD according to ISPOCD criteria after three months was the defined endpoint of the BioCog study and assessed in the designated POCDr package for R (Moller et al., 1998; Spies et al., 2021). Since the incidence of POCD in the sample presented here was very low (Lachmann et al. 2019), we decided to analyze the association of SWI hypointensities with a continuously scaled parameter for global cognitive function. Therefore, we calculated the global cognitive component ‘g’ to assess cognitive decline after surgery as described in previous publications (Feinkohl et al., 2019; Feinkohl, Janke, et al., 2020; Lammers et al., 2018).

At baseline assessment before surgery, all participants underwent a comprehensive computerized neuropsychological test battery (CANTAB, Cambridge Cognition Ltd., UK) and additional tests (Trail-Making Test, TMT-B, and the Grooved Pegboard Test, GPT). The CANTAB test battery comprised the Simple Reaction Task (SRT), Paired Associate Learning test (PAL), Verbal Recognition Memory (VRM) and the Simple Span test (SSP). The tests have been described elsewhere in detail (Lammers et al., 2018). Testing was performed by trained staff based on a standard operating procedure which was consented to with two neuropsychologists.

#### 2.3.1 Calculation of the global component ‘g’

Here, we calculated a modified version of the global cognitive component ‘g’ which had already been described in earlier publications from the BioCog study (Feinkohl et al., 2019; Feinkohl, Janke, et al., 2020; Lammers et al., 2018): ‘g’ is derived as the first component of a principal component analysis (PCA) of multiple cognitive tests.

Since the derivation of ‘g’ requires a complete neuropsychological assessment, we applied multiple imputations using chained equations to replace missing data in patients who began but did not complete the assessment. Missing cognitive data at baseline were imputed from available data at baseline, and missing postoperative cognitive data were imputed from available postoperative data. Results are based on 1000 imputations using predictive mean modelling. As described previously, PCA of performance in TMT-B, GPT, SRT, PAL and VRM immediate free recall was used to derive ‘g’ from the first principal component. TMT-B, GPT and SRT were log-transformed and reversed, and all variables were centred to the mean and normalized prior to PCA.

To calculate postoperative ‘g’, postoperative cognitive test parameters underwent the same transformations using centering and normalization parameters from preoperative data. We applied the rotation matrix derived from PCA of preoperative data to the postoperative cognitive data to calculate postoperative ‘g’.

### 2.4 Neuroimaging

#### 2.4.1 MRI acquisition

As a subproject of the BioCog study, a SWI sequence was added to the MRI protocol of the ongoing study. All patients underwent a standardized MRI protocol including a 3D susceptibility weighted gradient echo sequence (SWI: voxel size: 0.7×0.6×1.2mm^3^, field of view: 230×180mm^2^ in 120 transversal slices, TR=28ms, TE=20ms, 15° flip angle) designed to detect cerebral microbleeds, and a 3D T1 magnetization-prepared rapid acquisition gradient echo sequence (MPRAGE in 192 sagittal slices, FOV=256×256mm^2^, 1mm^3^ isotropic voxels, TR=2500ms, TE=4.77ms, 7° flip angle, parallel imaging with generalized autocalibrating partially parallel acquisitions using 24 reference lines, acceleration factor R=2). Of note, phase images from SWI were not saved, and hence, superior approaches to measure iron deposition (e.g., quantitative susceptibility mapping) were not performed. Data were acquired on one single 3T Magnetom Trio RIM MR scanner (Siemens) equipped with a 32-channel head coil at the Berlin Center for Advanced Neuroimaging (BCAN).

#### 2.4.2 Rationale of SWI-derived biomarkers

Hypointensities in SWI may originate from various sources of dia- and paramagnetic substances, such as blood products (e.g., hemosiderin) as post-hemorrhagic remnants, iron or calcium content and intravenous deoxyhemoglobin (Haller et al., 2021). Given that highly specific neuroimaging approaches for detection of cerebral iron deposition are available, few studies have used SWI to assess hypointensity as a surrogate parameter for iron and calcifications. Apart from semiquantitative rating methods, previous studies applied various intensity-derived metrics to measure brain mineralization in SWI: Harder and Schneider reported intensity per area as atrophy-adjusted parameters of mineralization (Harder et al., 2008; Schneider et al., 2016), whereas Gupta and Meijer used CSF-normalized measures of intensity to adjust for inconsistencies in a reference standard (Gupta et al., 2010; Meijer et al., 2015).

In this study, we applied a similar normalization procedure as the groups of Gupta and Meijer (Gupta et al., 2010; Meijer et al., 2015), i.e., relative intensity in the regions of interest compared to mean intensity of the whole brain white matter, as this parameter best reflected patterns of age-associated iron deposition (see supplementary figure S1). To address concerns of confounding by regional atrophy posed in the works by Harder and Schneider, ROI volumes were included as covariates in the analyses.

#### 2.4.3 Analysis of susceptibility-weighted images

Mean SWI intensities in each ROI (basal ganglia, hippocampus and pBFCS) were calculated following creation of binarized ROI maps from MPRAGE images using standard atlases coregistered to each patient scan. SPM12 (The Wellcome Centre for Human Neuroimaging, UCL Queen Square Institute of Neurology, London, UK, RRID: SCR_007037, http://www.fil.ion.ucl.ac.uk/spm/software/spm12/) in a MATLAB environment (The Mathworks. Inc. Natick. MA, RRID: SCR_001622), the log_roi_batch extension by Adrian Imfeld (http://www.aimfeld.ch/neurotools/neurotools.html) and FSLeyes (FMRIB, Analysis Group, Oxford, UK, RRID: SCR_002823, https://fsl.fmrib.ox.ac.uk/) were used for all MRI processing steps.

FSLeyes was used to create binary masks from the Harvard-Oxford subcortical (basal ganglia) and cortical probabilistic atlases. For the basal ganglia mask, we combined the 50% probability masks of the caudate nucleus, pallidum, putamen, and nucleus accumbens. For the hippocampus, 50% probability masks of the cornu ammonis, dentate gyrus, entorhinal cortex and subiculum were combined. The binary atlas of the BFCS had been described and provided by Laszlo Zaborszky and was already used in previous works (Lammers et al., 2018; Zaborszky et al., 2008). The pBFCS here refers to the combined regions of Ch4 and Ch4p.

SWI were coregistered to MPRAGE images of each patient. Grey and white matter as well as cerebrospinal fluid were parcellated from MPRAGE scans using the SPM12 segmentation routine and transformed to DARTEL (Diffeomorphic Anatomical Registration using Exponentiated Lie algebra) space. The segmented patient data were mapped onto a common template using the DARTEL flow fields implemented in SPM12, as described earlier (Heinrich et al., 2020; Lammers et al., 2018).

Atlas reference regions (MNI152 for hippocampus and basal ganglia, Colin27 for the pBFCS) were coregistered to a template. Using the SPM12 Deformations tool, a composition of the resulting DARTEL flow fields were applied to the binary ROI masks of the basal ganglia, the hippocampus and the pBFCS, resulting in individually labeled anatomical patient MRI data. Labeling of the white matter for normalization was achieved by binarizing the white matter probability maps generated by SPM at a threshold of 0.6. Mean signal intensity and volume of the ROI were derived using the log_roi_batch extension for SPM.

Finally, relative SWI intensities were calculated as the ratio of mean ROI SWI intensity and the mean white matter SWI intensity.

### 2.5 Statistical analysis

#### 2.5.1 Preoperative neuroimaging

We analyzed the association of relative SWI intensities in the basal ganglia, hippocampus and pBFCS and POD using generalized linear models with logit link function and assuming binomial error distribution (logistic regression). Analyses were conducted twice, with the first model including relative SWI intensity and duration of surgery as independent variables (reduced models), and the second model additionally including the patient characteristics age, sex, baseline MMSE and ROI volume as independent variables (extended models).

To study associations of relative SWI hypointensities in the three ROIs with postoperative cognitive decline after three months, we used a general linear model treating postoperative global cognitive function ‘g’ as the dependent variable and including preoperative ‘g’, preoperative relative SWI intensity and surgery duration as independent variables (reduced models). Preoperative ‘g’ was included in the model with the intention to assess associations between preoperative hypointensities and postoperative cognitive decline rather than absolute postoperative cognitive function. We repeated the analysis with adjustment for age, sex, and ROI volume (extended models).

#### 2.5.2 Postoperative neuroimaging

Postoperative cognitive decline was analyzed in a general linear model treating the global cognitive component ‘g’ as the dependent variable. The reduced model included perioperative change in relative SWI intensities, preoperative ‘g’ and surgery duration as independent variables. The extended models additionally included age, sex, and postoperative ROI volume. The perioperative change in relative SWI intensity was calculated as the residual of postoperative relative SWI intensity after regression on preoperative relative SWI intensity.

Since the number of patients who experienced POD with postoperative SWI data was too low for a reliable analysis, these analyses are only available as supplementary material.

#### 2.5.3 Post-hoc analysis of pBFCS subregions

In a post-hoc analysis, we studied the associations of two subregions in the pBFCS (Ch4 and Ch4p) with POD and postoperative cognitive function. Based on our observations, post-hoc analyses were restricted to associations of relative preoperative hypointensity of Ch4 and Ch4p with POD as well as associations of perioperative changes in relative hypointensity of Ch4 and Ch4p with postoperative cognitive functions. Reduced and extended models included the same additional variables as described above.

We report effect sizes as regression coefficients with confidence intervals and model degrees of freedom (df). For linear regression models, we calculated adjusted R^2^ for the whole model and partial R^2^ for the SWI intensity measures. Additionally, we report odds ratios (OR) for logistic regression which have been normalized to a change in relative intensity by 0.1 (denoted as OR _0.1_) for interpretability, which approximately corresponds to a change by two standard deviations.

To account for low statistical power due to the small sample size, we decided to make prior assumptions about the direction of the association and calculate one-sided p-values. We expected lower (or a decrease in, respectively) relative SWI intensities to be associated with POD and lower postoperative cognitive function ‘g’. The level of significance was set at p<0.05 (one-sided) without adjustment for multiple testing and 90% confidence intervals were calculated. Results may therefore still be considered exploratory. Statistical analysis was conducted in R version 4.2.1 (Funny-Looking Kid, RRID: SCR_001905) with additional use of the car, factoextra, questionr, sensemakr and mice packages for analyses as well as ggplot2, jpeg, magick and cowplot for creation of figures.

## 3 Results

### 3.1 Sample

933 patients were enrolled in the whole BioCog study between 2014/11 and 2017/04. The SWI subproject was initiated 2016/04. 65 patients with preoperative SWI data were included in the analysis. 14/65 (22%) of the patients experienced POD and 40/65 (62%) returned for neuropsychological testing after three months. Overall POD rates were comparable to other analyses which used data from the BioCog cohort (i.e., 20% (Heinrich et al., 2021) and 25% (Lammers-Lietz et al., 2022)) as well as other studies conducted in our center (i.e., 16% (Spies et al., 2021) and 31% (van Norden et al., 2021)).

Since SWI was added to the MRI protocol during enrollment of patients, some patients underwent SWI at the follow-up session, but not before surgery. Postoperative SWI data were available for 54 patients in total, of whom only 34 patients had pre- and postoperative SWI. Among these, 3/34 (9%) experienced POD and 33/34 (97%) had two neuropsychological assessments before and three months after surgery. Tables 1 and 2 describe the preoperative (*N*=65) and longitudinal (*N*=33) samples. Patients flow charts are given in the supplementary material (figure S2).

**Table 1.** Demographic and clinical characteristics of the study sample with preoperative SWI data (N=65)

|  |  | N | % |
| --- | --- | --- | --- |
| **Women** |  | 35 | 54 |
| **ISCED** | **Level 1-2** | 5 | 9 |
|  | **Level 3-4** | 27 | 47 |
|  | **Level 5-6** | 25 | 44 |
| **Intrathoracic, -abdominal or -pelvic surgery** |  | 21 | 32 |
| **ASA Physical Status** | **I** | 1 | 2 |
|  | **II** | 41 | 63 |
|  | **III** | 23 | 35 |
| **(Pre-)Frailty** |  | 28 | 44 |
| **Hazardous alcohol consumption (AUDIT)** |  | 4 | 6 |
| **Anesthesia** | **Regional** | 2 | 3 |
|  | **General + regional** | 23 | 35 |
| **ICU admission** |  | 11 | 17 |
| **Postoperative pain<sup>a</sup>** |  | 27 | 44 |
| **Postoperative complication** |  | 31 | 48 |
| **POD** |  | 14 | 22 |
| **90d survival** |  | 2 | 3 |
|  | Median | Interquartile range | Min.-max. range |
| **Age (y)** | 72 | 68-75 | 65-87 |
| **CCI (p)** | 1 | 0-2 | 0-5 |
| **Baseline MMSE (p)** | 29 | 28-30 | 24-30 |
| **Baseline 'g'** | 0.33 | -0.88-1.53 | -2.45-4.15 |
| **Postoperative 'g'** | 0.58 | -0.49-1.66 | -3.96-3.60 |
| **Surgery duration (min)** | 105 | 70-197 | 3-495 |
| **Days in hospital** | 7 | 5-10 | 2-67 |
| <sup>a</sup> Positive screening from a compound assessment including the Non-visual Rating Scale (NRS), Behavioral Pain Scale (BPS and BPS-NI) and Critical Pain Observation Tool (CPOT) during the first seven postoperative days<br>Abbreviations: ASA, American Society of Anesthesiologists; AUDIT, Alcohol Use Disorder Identification Test; CCI, Charlson Comorbidity Index; d, days; ISCED, International Standard Classification of Education; Max., Maximum; min, minutes; Min., Minimum; MMSE, Mini-mental Status Examination; p, points; y, years; |  |  |  |

**Table 2.** Demographic and clinical characteristics of the study sample with longitudinal SWI and neuropsychological data (N=33)

|  |  | N | % |
| --- | --- | --- | --- |
| **Women** |  | 17 | 52 |
| **ISCED** | **Level 1-2** | 2 | 7 |
|  | **Level 3-4** | 13 | 43 |
|  | **Level 5-6** | 15 | 50 |
| **Intrathoracic, -abdominal or -pelvic surgery** |  | 5 | 15 |
| **ASA Physical Status** | **I** | 1 | 3 |
|  | **II** | 21 | 64 |
|  | **III** | 11 | 33 |
| **(Pre-)Frailty** |  | 13 | 41 |
| **Hazardous alcohol consumption (AUDIT)** |  | 1 | 3 |
| **Anesthesia** | **Regional** | 1 | 3 |
|  | **General + regional** | 11 | 33 |
| **ICU admission** |  | 1 | 3 |
| **Postoperative pain** |  | 12 | 61 |
| **Postoperative complication** |  | 14 | 42 |
| **POD** |  | 3 | 10 |
|  | Median | Interquartile range | Min.-max. range |
| **Age (y)** | 72 | 67-75 | 65-86 |
| **CCI (p)** | 0 | 0-2 | 0-5 |
| **Baseline MMSE (p)** | 28 | 28-30 | 25-30 |
| **Baseline 'g'** | 0.02 | -0.90-1.15 | -2.45-4.15 |
| **Postoperative 'g'** | 0.64 | -0.53-1.52 | -3.96-3.42 |
| **Surgery duration (min)** | 138 | 107-182 | 56-404 |
| **Days in hospital** | 7 | 4-8 | 2-12 |
| Abbreviations: see table 1. |  |  |  |

### 3.2 Associations of preoperative relative SWI hypointensities with POD

Figure 1 displays relative SWI intensity for all three regions of interest in all patients separated by POD in *N*=65 patients. Adjusted for duration of surgery, relative SWI hypointensities in the basal ganglia and the pBFCS, but not in the hippocampus, were associated with increased risk for POD. After additional adjustment for age, sex, baseline MMSE and ROI volume, the association persisted for the pBFCS, but neither for basal ganglia nor the hippocampus. See figure 1 for details on the statistical results.

**Figure 1.**
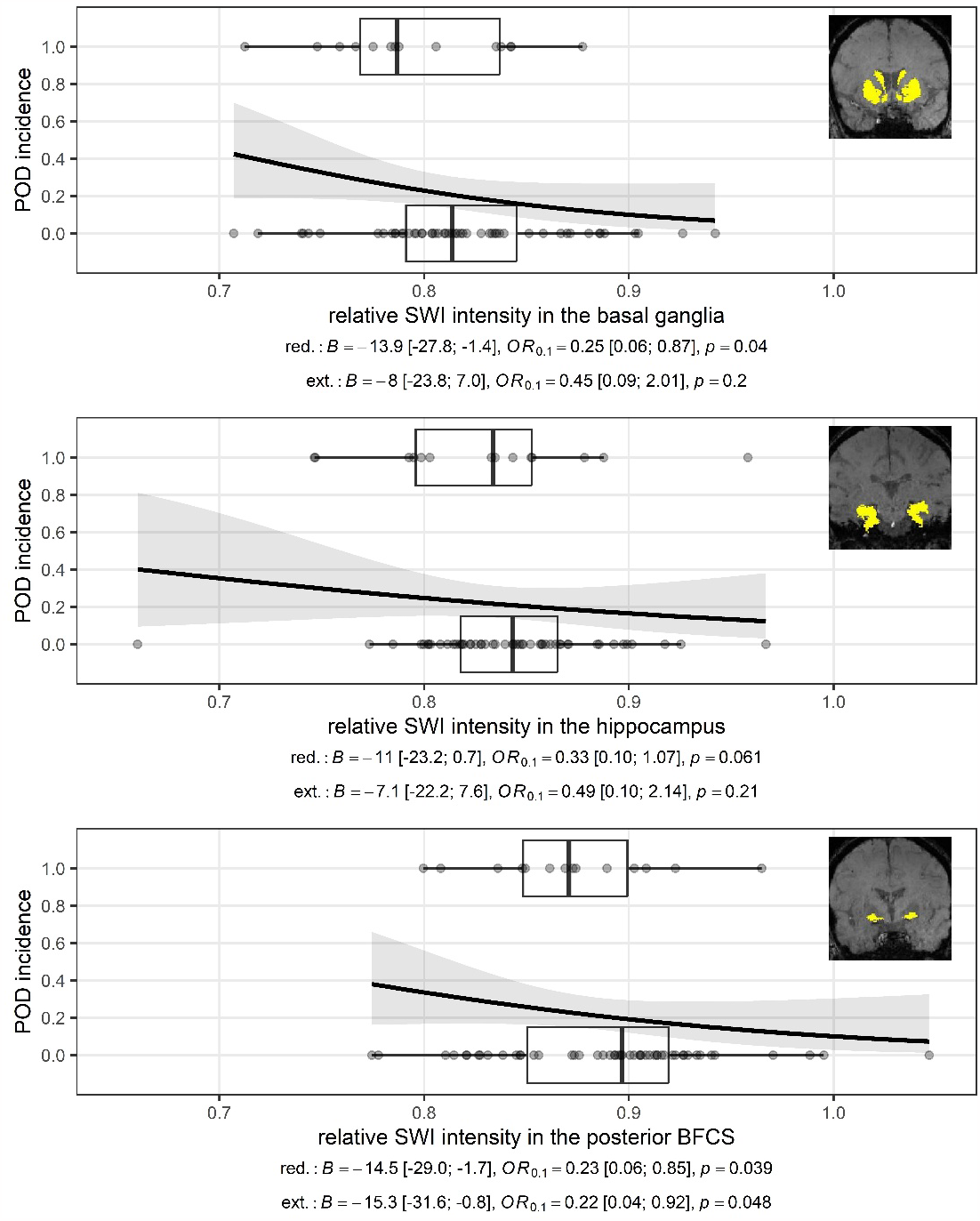
displays the association of relative SWI intensity (x-axis) in three regions of interest (ROI) with POD (y=axis) in N=65 patients as dot plots. Horizontal boxplots summarize the distribution of SWI relative intensity values for patients who experienced POD (top in each plot, POD incidence=1) and those who did not (bottom in each plot, POD incidence=0). The line shows the conditional POD incidence for each relative SWI intensity value with 90% CI (shaded area) based on a simple logistic regression. Lower values for the relative SWI intensity indicate that the ROI appears darker in the MRI, suggesting higher mineralization. Intensities must be interpreted with reference to the mean intensity of the white matter, i.e., a value of 1 means equal signal intensity of the ROI and white matter, whereas values <1 indicate hypointense appearance of the ROI compared to the white matter. Inlays display the regions of interest. The pBFCS here refers to the combined regions of Ch4 and Ch4p in Zaborzsky’s modification of Mesulam’s nomenclature of the cholinergic system (Zaborzsky et al., 2008). Details of the statistical models (regression coefficients B, odds ratios for 0.1 change in relative SWI intensity with 90% confidence intervals and one-sided p-value are given in the plot captions. Degrees of freedom are 2/62 for all reduced, and 6/58 for all extended models. Abbreviations: B: logistic regression coefficient; BFCS: basal forebrain cholinergic system; ext.: extended model results; OR_0.1_: odds ratio for 0.1 unit change in relative intensity; p: p-value; POD, postoperative delirium; SWI: susceptibility-weighted imaging; red.: reduced model results

### 3.3 Associations of preoperative relative SWI hypointensities with postoperative decline global cognitive performance ‘g’

In *N*=40 patients with preoperative SWI data and longitudinal neuropsychological testing, neither relative hypointensities in the basal ganglia (*B*=3.64 [-1.71; 9.00], *R*^2^=0.74, partial *R*^2^=0.035, *p*=0.13), nor hippocampus (*B*=0.86 [-5.97; 7.70], *R*^2^=0.73, partial *R*^2^=0.001, *p*=0.42), nor pBFCS (*B*=-0.71 [-5.81; 4.40], *p*=0.59, *R*^2^=0.73, partial *R*^2^=0.002) were associated with postoperative ‘g’ after adjustment for surgery duration and preoperative ‘g’ (*df*=3/36 for all reduced models). Results were not changed after adjustment for age, sex, and ROI volume (basal ganglia: *B*=2.94 [-2.65; 8.52], *R*^2^=0.75, partial *R*^2^=0.023, *p*=0.45; hippocampus: *B*=-1.71 [-8.94; 5.52], *R*^2^=0.73, partial *R*^2^=0.005, *p*=0.65; posterior BFCS: *B*=-0.61 [-5.67; 4.44], *p*=0.58, *R*^2^=0.74, partial *R*^2^=0.001; *df*=6/33 for all extended models).

### 3.4 Associations of perioperative changes in relative SWI hypointensities with postoperative decline global cognitive performance ‘g’

pBFCS intensity changes in *N*=33 patients with pre- and postoperative SWI data and neurocognitive testing at follow-up were significantly positively associated baseline-adjusted postoperative ‘g’ in the reduced model, and in the extended model on a trend level, i.e., a further postoperative decrease of relative SWI intensity was associated with lower postoperative global cognitive function. Significant associations were neither observed in the hippocampus nor the basal ganglia (see figure 2 for details on the statistical results).

**Figure 2.**
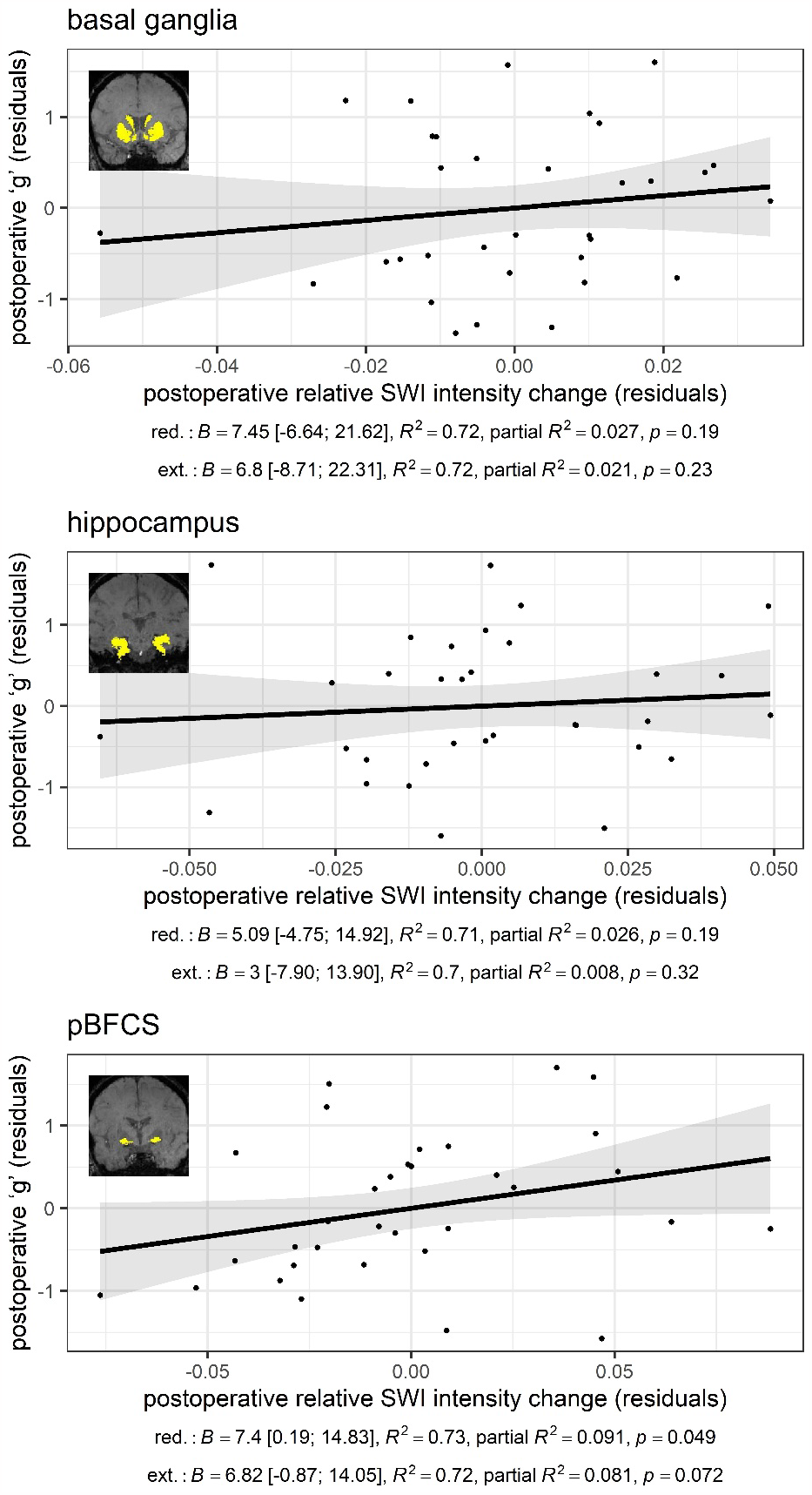
displays associations of perioperative relative SWI intensity change in the three regions of interest with postoperative global cognitive function ‘g’ at follow-up three months after surgery in N=33 patients. The graphs are partial regression plots of the extended models adjusted for preoperative ‘g’, surgery duration, age, sex and postoperative ROI volume. Inlays display the regions of interest. The captions give regression coefficients B with 90% confidence intervals, adjusted R^2^ for the whole model and partial R^2^ for the SWI intensity measure and p-values for the reduced and extended models. Degrees of freedom are 3/29 and 6/26 for reduced and extended models, respectively. Abbreviations: red.: reduced; ext.: extended

### 3.5 Post-hoc analyses of pBFCS subregions

Relative preoperative SWI hypointensities in Ch4 were significantly associated with POD in the reduced model and at a trend level in the extended model, but no significant association was observed for Ch4p. We observed significant associations of postoperative decrease in relative SWI intensities of Ch4p with postoperative ‘g’, which were independent of age, sex, preoperative ‘g’ and ROI volume, but not Ch4. Hence, postoperative longitudinal decline in cognition was associated with progressing postoperative hypointensities in region Ch4p. Results from the post-hoc analysis including statistical details are displayed in figure 3.

**Figure 3.**
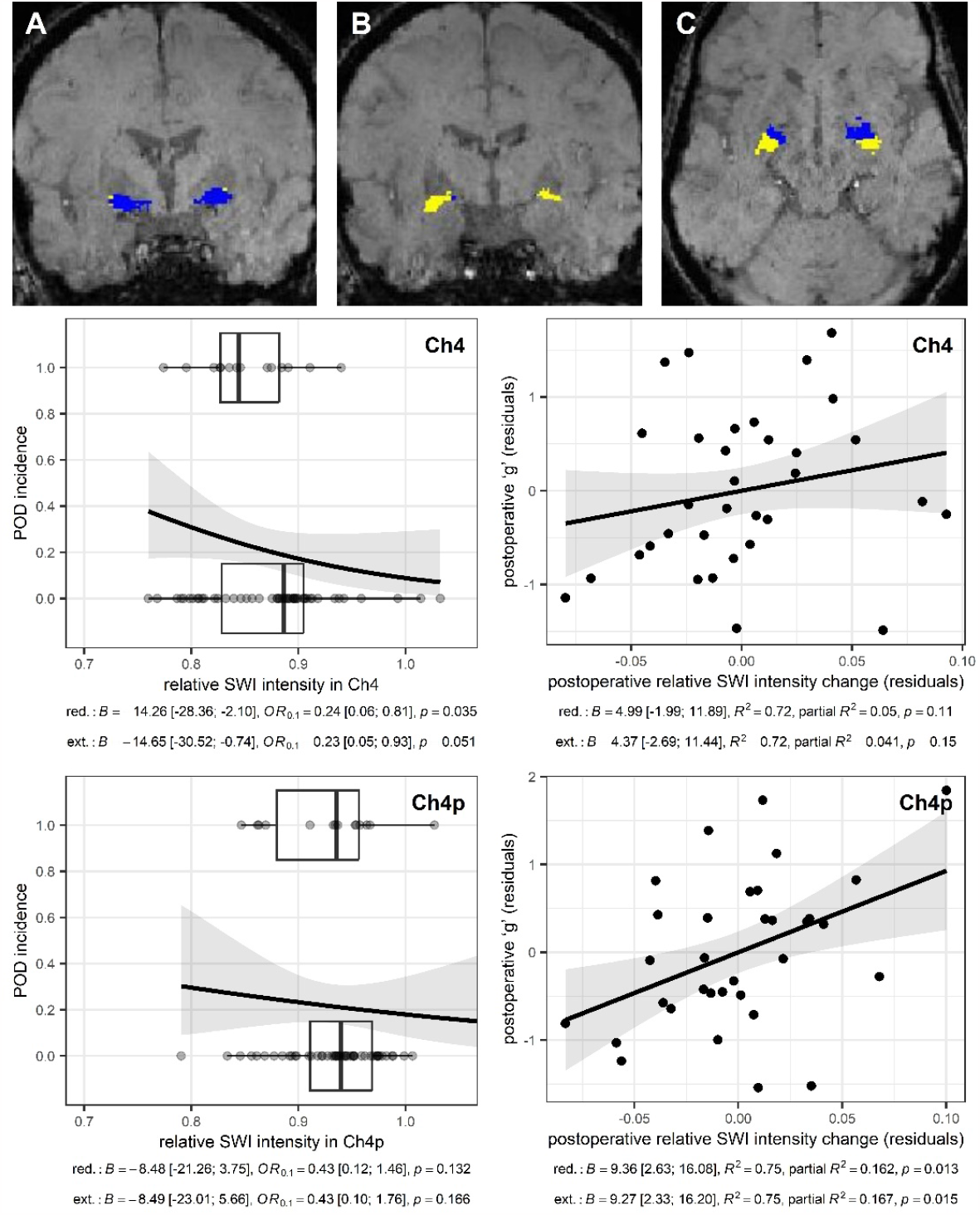
displays the two subregions of the pBFCS (top row) and the results of the post-hoc analysis (middle and bottom row). In the top row, Ch4 (blue) and Ch4p (yellow) are depicted in coronal (A, B) and one axial slice (C). Associations of regions Ch4 (middle) and Ch4p (bottom) with POD (left, N=65) and postoperative cognitive function (right, N=33). Degrees of freedom are 2/62 and 6/58 for reduced and extended models of POD, respectively, as well as 3/29 and 6/26 for models of ‘g.’ For a detailed explanation of the figures, see also figures 1 and 2.

## 4 Discussion

Here, we studied the association of SWI hypointensities in the basal ganglia, the hippocampus and the pBFCS with POD and postoperative cognitive decline.

We observed an association of preoperative hypointensity of the pBFCS and basal ganglia with POD, but only in the pBFCS, this association was found to be independent of confounders such as age, MMSE, sex and region volume. A post-hoc analysis suggested that relative hypointensity in the rostral pBFCS, region Ch4, might be more relevant for POD than Ch4p. Ch4 has been described as the main source of cholinergic innervation to associative and sensory frontoparietal cortical areas (Mesulam et al., 1983). Although iron deposition in the BFCS has rarely been investigated, and existing studies did not report an association with neurodegenerative disease (Gu et al., 1998), diffusion tensor imaging studies reported brain tissue alterations in the BFCS as a predisposing factor for POD (Cavallari et al., 2016). In accordance, studies reported altered connectivity of the cholinergic system, especially region Ch4, in Lewy body dementia with hallucinations (Hepp et al., 2017; Mehraram et al., 2022).

In contrast, preoperative hypointensity was not associated with decline in global cognitive function three months after surgery. However, a postoperative decrease of pBFCS intensity was associated with postoperative decline in cognitive function in the reduced model, and in the extended model at a trend level. In the post-hoc analysis, we found that this association was driven by a significant association of cognitive decline and relative SWI hypointensity in the most posterior part of the pBFCS (Ch4p), which was independent of age, sex, and region volume in the extended model. Hence, SWI hypointensity as a surrogate parameter for neurodegeneration seems to contribute differentially to POD and postoperative cognitive decline. Whereas it is a predisposing factor for POD, postoperative progress of mineral and iron deposition in the pBFCS reflect ongoing neurodegenerative processes related to postoperative cognitive decline. The post-hoc analysis revealed an association with SWI intensity changes in the Ch4p, which is assumed to provide cholinergic innervation of the temporal lobe, rather than the whole pBFCS (Mesulam et al., 1983). Previous imaging studies reported early atrophy of Ch4p in AD (Kilimann et al., 2017). Of note, cerebral iron accumulation has also been reported to be associated with both amyloid plaques and tau fibril aggregates in AD, and hence, our findings may point to common pathways in postoperative cognitive decline and AD (Collingwood et al., 2005; Spotorno et al., 2020).

Interestingly, we found no independent association of hypointensity in the basal ganglia or the hippocampus, although these regions are commonly reported to be vulnerable to mineralization and calcification in any of the extended models (Acosta-Cabronero et al., 2016; Betts et al., 2016; Burgetova et al., 2021; Haacke et al., 2005). This might be due to methodological aspects, since the relative hypointensity measure was chosen based on this association with age, and hence, distinction between effects of age and hypointensity on postoperative neurocognitive disorders are difficult to make. On the other hand, the pBFCS may be a particularly vulnerable region for mineralization and calcification and may have a critical role for the pathogenesis of POD and postoperative cognitive decline: In fact, the Successful Aging after Elective Surgery study group reported white matter alterations in the BFCS as a risk factor for POD (Cavallari et al., 2016) and one anecdotal case report on donepezil-responsive delirium due to severe lesioning of the basal forebrain after surgical removal of a craniopharyngeoma exist (Kobayashi et al., 2004). Various studies reported increased sensitivity to anesthetics in animals with BFCS lesions (Leung & Luo, 2021; Leung et al., 2014) and it has been shown that animals were more susceptible to inflammation-induced cognitive deterioration after lesioning the BFCS, suggesting a role in septic encephalopathy (Field et al., 2012). BFCS integrity was an essential mediator for the therapeutic effect of vagal nerve stimulation in a rat model of POCD, suggesting that cortical acetylcholine released from the BFCS may inhibit overshooting neuroinflammation after anesthesia (Zhou et al., 2023).

The interpretation of hypointensity in SWI is difficult, as it may originate from cerebral bleeding, iron or calcium content and even intravenous deoxyhemoglobin (Haller et al., 2021). Calcifications are common findings in cerebral imaging of otherwise healthy aging patients, although they may also originate from various diseases and may be associated with neuropsychiatric symptoms (Harrington et al., 1981; Saade et al., 2019). A recent review on neuroimaging findings in COVID19 reported microhemorrhages to be a common finding in COVID19-associated encephalopathy (Ghaderi et al., 2023). Microbleeds in the pBFCS have not been assessed in this study, however, in the previous analysis of microbleeds conducted in the same sample, there was no significant association of microbleeds with POD or POCD, suggesting that posthemorrhagic remnants and the associated increase in neurovascular risk are not the major mediator in the association of SWI hypointensities with POD or postoperative cognitive decline. Furthermore, the prevalence of deep microbleeds possibly affecting our regions of interest was very low in the investigated sample (4 out of 65 patients) (Lachmann et al., 2019).

This is the first study on brain mineralization in postoperative neurocognitive disorders. Since we tested our hypothesis post-hoc in an available dataset, the study has certain methodological shortcomings. First, SWI was added to the MRI protocol of an ongoing study without prior statistical planning. Hence, the statistical power is low, and all results need to be considered exploratory, warranting larger validation studies. Age is a relevant cofounder in our study, especially since the normalization procedure was chosen based on the correlation of normalized SWI intensities with age. Of note, relative hypointensity of basal ganglia was no longer significant after adjustment for additional variables including age. Hence, results from the reduced models may be confounded by age, whereas additional inclusion of age in the extended models may result in overcorrection of the model leading to false-negative results. Furthermore, the MRI protocol was designed for microbleeds assessments rather than measurement of iron or calcium deposition, whereas advanced neuroimaging methods such as quantitative susceptibility mapping, R2* relaxometry and biophysical modelling approaches have been developed to quantify cerebral iron deposition (Drori et al., 2022; Haacke et al., 2005). Hence, our results are somewhat indiscriminate to the etiology of hypointensities, as outlined above and additional studies with MRI protocols designed to measure brain tissue composition are needed (Chawla et al., 2018; Chawla et al., 2016; Cooper et al., 2020; Wieland et al., 2021). Results from these studies could then justify trials, i.e., on iron chelation therapy in the perioperative setting, as preliminary studies suggest efficacy in several neurodegenerative diseases, including Parkinson’s disease and Alzheimer’s dementia (Nunez & Chana-Cuevas, 2018).

## 5 Conclusion

We present the first, although highly exploratory study on brain tissue mineralization in postoperative neurocognitive disorders. Our results suggest that brain mineralization, particularly in the cholinergic system, is a possible contributor to POD and postoperative cognitive decline. Whereas preoperative relative SWI hypointensities seem to be related to patients predisposed to POD, tissue changes leading to lasting cognitive decline in surgical patients seem to occur in the postoperative period. Although warranting further studies, our results imply there could be a prognostic benefit in added treatment options, i.e., chelation therapy or calcium channel blockade, for patients with or at risk for postoperative neurocognitive disorders.

## Data Availability

Data sharing is not applicable to this article as no new data were created or analyzed in this study. Data from the BioCog study are not publicly available due to constraints imposed in the consent forms. An anonymized version is available from the authors on reasonable request.

## 6 Acknowledgements

## 6.2 Competing interests

Florian Lammers-Lietz, MD, received personal fees from PI Health Solutions GmbH during the conduct of the study.

Friedrich Borchers, MD: none

Stefan Hetzer, PhD: none

Insa Feinkohl, PhD: none

Cicek Kanar, BSc: none

Gunnar Lachmann, MD, PhD: none

Claudia Paarmann-Chien, PhD, received research funding from Novartis and Alexion, unrelated to this study and is a Standing Committee on Science Member for the Canadian Institutes of Health Research (CIHR).

Claudia Spies, MD, PhD, received grants from the European Commission during the conduct of the study. During the past 36 months, Prof. Spies received grants from Deutsche Forschungsgemeinschaft, Deutsches Zentrum für Luft- und Raumfahrt e.V., Einstein Stiftung Berlin, Federal Joint Committee (GBA Innovationsfond), inner university grants, Projektträger im DLR, Stifterverband, Federal Ministry for Economic Affairs and Climate Action, payments by the Georg Thieme Verlag, sponsoring from Dr. F. Köhler Chemie GmbH, Sintetica GmbH, Federal Joint Committee (GBA Innovationsfond), Max-Planck-Gesellschaft zur Förderung der Wissenschaften e.V., Stifterverband für die deutsche Wissenschaft, Philipps Electronics Nederland BV, Federal Ministry of Education and Research, Robert-Koch-Institut and the European Commission. Prof. Spies is involved in patents 15753 627.7 (issued), PCT/EP 2015/067731 (issued), 3 174 588 (issued), 10 2014 215 211.9, 10 2018 114 364.8, 10 2018 110 275.5, 50 2015 010 534.8, 50 2015 010 347.7, 10 2014 215 212.7. She participates in or is member of the Association of the Scientific Medical Societies in Germany (AWMF), Deutsche Forschungsgemeinschaft and the German National Academy of Sciences (Leopoldina) without receiving payments.

Georg Winterer, MD, PhD: grants from the European Commission during the conduct of the study. CEO of PharmaImage Biomarker Solutions GmbH Berlin (Germany) and President of its subsidiary Pharmaimage Biomarkers Incl. (Cambridge, MA, USA) and PI Health Solutions GmbH Berlin (Germany). Grants from the Deutsche Forschungsgemeinschaft/German Research Society and from the German Ministry of Health.

Laszlo Zaborszky, PhD: none

Norman Zacharias, PhD: none

Friedemann Paul, MD, PhD: none

## 6.3 Authors’ contributions

Florian Lammers-Lietz, MD: Conceptualization, Methodology, Formal analysis, Investigation, Data Curation, Writing – Original Draft & Editing, Visualization; Friedrich Borchers, MD: Methodology, Formal analysis, Investigation, Data Curation; Insa Feinkohl, PhD: Methodology; Stefan Hetzer, PhD: Methodology; Cicek Kanar, BSc: Formal Analysis; Gunnar Lachmann: Methodology, Investigation; Claudia Spies, MD, PhD: Conceptualization, Resources, Supervision, Project Administration, Funding acquisition; Claudia Chien: Methodology, Writing – Original Draft & Editing; Claudia Spies: Conceptualization, Resources, Supervision, Project Administration, Funding acquisition, Data Curation; Georg Winterer, MD, PhD: Conceptualization, Resources, Project Administration, Data Curation, Funding acquisition; Laszlo Zaborszky: Resources; Norman Zacharias, PhD: Methodology, Investigation, Formal Analysis; Friedemann Paul, MD, PhD: Conceptualization, Methodology, Investigation, Writing – Original Draft & Editing, Supervision. All authors have reviewed and approved the final version of the manuscript for publication.

## 6.4 Funding

The BioCog Project was funded by the European Union Seventh Framework Program [FP7/2007-2013] under grant agreement n° 602461. Gunnar Lachmann was participant in the BIH Charité Clinician Scientist Program funded by the Charité-Universitätsmedizin Berlin, and the BIH at Charité.

## 6.5 Last notes

We would like to express our gratitude to the whole BioCog study team, including the study team at the UMC Utrecht and Dr. Konrad Neumann (Institut für Biometrie und Klinische Epidemiologie at Charité-Universitätsmedizin Berlin) for his statistical advice. PI Health Solutions GmbH (https://pi-healthsolutions.com/) and the Pharmaimage team are acknowledged for their extensive contributions during the conduct of the BioCog study.

## Notes

### Clinical Protocols

https://classic.clinicaltrials.gov/ct2/show/NCT02265263

### Funding Statement

The BioCog Project was funded by the European Union Seventh Framework Program [FP7/2007-2013] under grant agreement no. 602461. Gunnar Lachmann was participant in the BIH Charite Clinician Scientist Program funded by the Charite-Universitaetsmedizin Berlin, and the BIH at Charite.

### Author Declarations

All study procedures were conducted in line with the declaration of Helsinki with approval by the local medical ethics committees of the study centres in Berlin, Germany (Charite-Universitaetsmedizin Berlin, EA2/092/14) and Utrecht, Netherlands (University Medical Center, 14-469).

