## supplementary for "Brain mineralization in postoperative delirium and cognitive decline"

### Supplementary material on the relative hypointensity measure

To assess the plausibility of various suggested susceptibility weighted image (SWI)-derived surrogate measures of brain mineralization, we compared the correlation of age with absolute mean SWI intensity and two relative SWI measures: the ratio of mean intensity in the region of interest (ROI) and mean white matter intensity as well as the ratio of mean ROI intensity and mean occipital cortex intensity. Mean white matter intensity was derived from the whole white matter tissue map (voxels with ≥0.6 probability), whereas mean occipital cortex intensity was derived from the 0.5 probability map of the occipital cortex of the Harvard-Oxford cortical atlas implementation in FSLeyes. The occipital cortex mask was chosen due to previous reports of relative low age-associated iron deposition in this cortical area (Acosta-Cabronero et al., 2016; Betts et al., 2016).

Pearson's correlation coefficients of age and SWI intensity in all three ROI was calculated for all three measures.

Supplementary figure S1 summarizes the results. In none of the three ROIs, absolute image intensity was correlated with age. We observed that in general, correlation with age increased after adjustment for cortical or white matter intensity by calculation of the ratio between image intensities in the ROI and cortex or white matter, respectively. Correlations were slightly stronger for the ratio in relation to white matter than to the occipital cortex, especially in regions with reported age-associated iron deposition. This is in line with previous reports which suggested lower general iron deposition in the white matter compared to grey matter (Haacke et al., 2005), as well as stronger age-related iron deposition in grey compared to white matter (Acosta-Cabronero et al., 2016). Independently of the region used for adjustment, we observed a stronger correlation of SWI intensity ratio with age in the basal ganglia compared to the hippocampus, and the lowest correlations for the pBFCS. This is in accordance with previous reports of strong age-related iron deposition in the basal ganglia and less prominent associations in the hippocampus, whereas no such findings have been reported for the BFCS (Acosta-Cabronero et al., 2016; Betts et al., 2016; Burgetova et al., 2021). Hence, our findings suggest that relative SWI hypointensities as reflected by the SWI ratio between the regions of interest assessed here is a probable surrogate parameter of (age-related) iron deposition. White matter should be preferred over grey matter as a control region for calculation of the control region, most likely due to its lower vulnerability for iron deposition in aging.

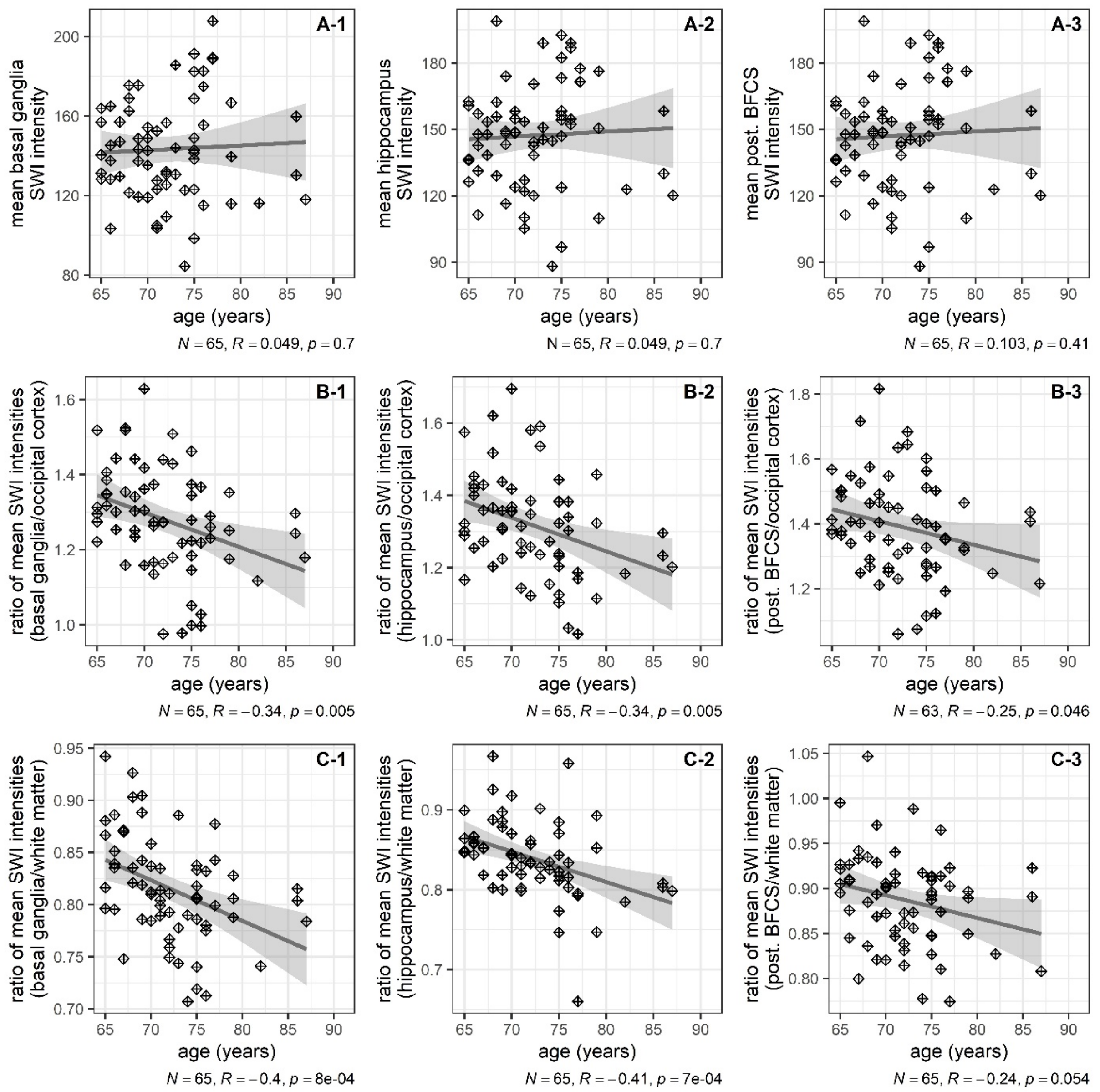

**Figure S1.** Correlations of three measures of hypointensity for three regions of interest with age as scatter plots and additional linear regression line with 95% confidence interval. Each column corresponds to one region of interest ROI: basal ganglia (left column, A-1, B-1 and C-1), hippocampus (middle column, A-2, B-2, C-2) and posterior BFCS, right column, A-3, B-3, and C-3). Age in years is given on the x-axis. In the top row (A-1 to A-3), absolute signal intensity is given, whereas lower rows show relative signal intensity as the ratio between the region of interest and the occipital cortex (middle row, B-1 to B-3) and the white matter (bottom row, C-1 to C-3). Annotations indicate sample size (*N*), Pearson's correlation coefficient (*R*) for the correlation between age and image intensity with *p*-value (two-tailed). Degrees of freedom are 1/63.

#### Supplementary material on the calculation of 'g'

The first principal component 'g' explained 40.4% of the total variance in the cognitive data, whereas the second to fifth component each explained ≤15.3%. Table S1 provides the factor loadings. 'g' was higher at baseline in patients who returned for testing after three months. Among patients with neuropsychological testing at follow up, postoperative 'g' was higher at follow-up three months compared to baseline (see table S2). Correlation between pre- and postoperative global cognitive performance was high (Pearson's $R$ [95% CI] =0.79 [0.76; 0.82], $p<0.001$).

**Table S1. Factor loadings of cognitive tests on preoperative global cognitive component 'g'.** Abbreviations: TMT-B, Trail-making test (time for completion, reversed and log-transformed), part B; GPT, Grooved pegboard test (time for completion, reversed and log-transformed); PAL, Paired associate learning test (first trial memory score); SSP, Simple span length (number of items); VRM, Verbal recognition memory (number of correctly remembered items at immediate recall); SRT, Simple reaction task (latency of correct responses, reversed and log-transformed)

| Cognitive test parameter | Loading on 'g' |
|---|---|
| **TMT-B** | 0.50 |
| **GPT** | 0.45 |
| **PAL** | 0.44 |
| **SSP** | 0.37 |
| **VRM** | 0.36 |
| **SRT** | 0.31 |

**Table S2. Distribution of 'g' at baseline and at follow-up three months after surgery.** Abbreviations: IQR, interquartile range; min., minimum; max., maximum

| | Median | IQR | Min.-max. range |
|---|---|---|---|
| **Preoperative 'g' in all patients with baseline testing (*N*=928)** | 0.05 | -1.04-1.09 | -5.06-4.15 |
| **Preoperative 'g' in all patients with baseline and postoperative testing (*N*=641)** | 0.31 | -0.74-1.30 | -4.48-4.15 |
| **Postoperative 'g' in all patients with baseline and postoperative testing (*N*=641)** | 0.59 | -0.47-1.60 | -4.86-4.81 |

#### Supplementary material: patient flow charts

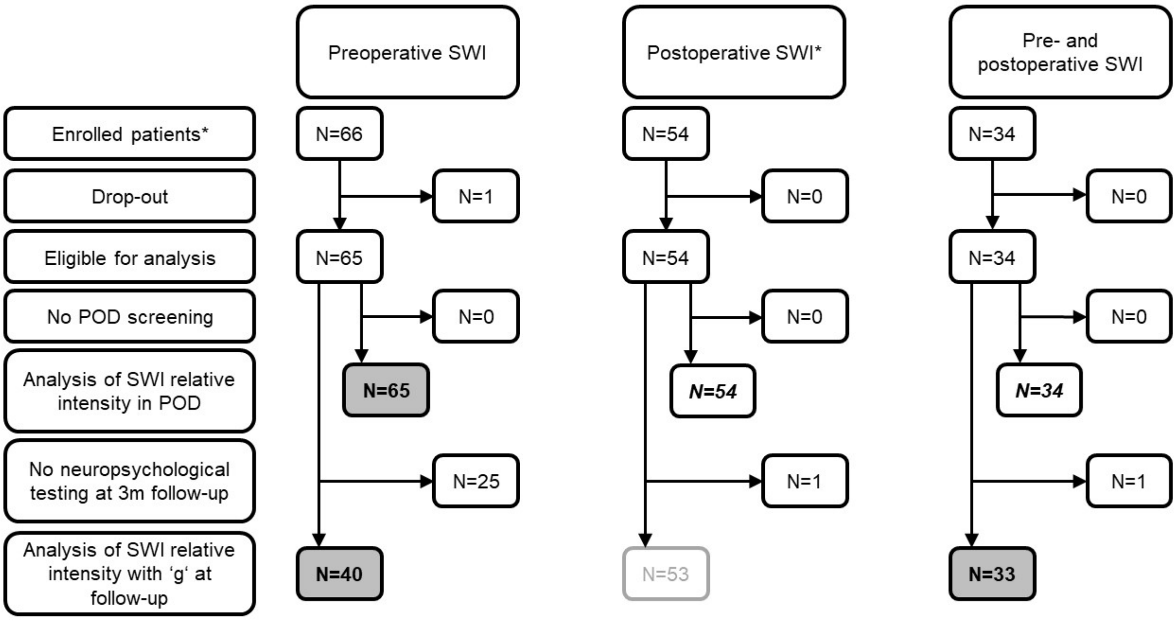

**Figure S2.** Patient flow charts. Grey shading and bold type setting indicate an endpoint analysis presented in the manuscript. Endpoints/samples in italic/bold type setting without shading indicate endpoint analyses only presented in the supplementary material for completeness since the number of cases with POD was too few.

*Since susceptibility-weighted imaging was added to the ongoing BioCog study as part of a subproject, SWI is available for only a subsample of all included patients (*N*=933 in the whole BioCog study) only. For the same reason, some patients underwent SWI at the follow-up assessment, but not at the preoperative neuroimaging session.

### Supplementary material to the analysis of postoperative MRI data

The number of patients with POD who had follow-up SWI data was generally too low for a comprehensive analysis. For completeness, the regression coefficients with 90% CI of two analyses using these data are given here, e.g., for use in power calculations.

**Table S3. Analysis of longitudinal SWI from 34 patients (N=3 with POD).** Effect sizes for the association of POD with postoperative relative SWI intensity (dependent variable) are given. Abbreviations: B, regression coefficient; CI, confidence interval

| | **Reduced model[a]** | | | **Extended model[b]** | | |
|---|---|---|---|---|---|---|
| **ROI** | *B* | *90% CI* | *Partial $R^2$* | *B* | *90% CI* | *Partial $R^2$* |
| **Basal ganglia** | 0.002 | -0.020-0.024 | <0.001 | 0.007 | -0.017-0.030 | 0.009 |
| **Hippocampus** | 0.001 | -0.029-0.033 | <0.001 | -0.002 | -0.033-0.029 | <0.001 |
| **pBFCS** | 0.013 | -0.027-0.055 | 0.010 | 0.012 | -0.032-0.056 | 0.008 |

[a] Linear regression adjusted for preoperative relative SWI intensity and duration of surgery (*df*=3/30)

[b] Linear regression adjusted for preoperative relative SWI intensity and duration of surgery, age, sex, preoperative MMSE and ROI volume (*df*=7/26)

**Table S4. Analysis of postoperative SWI from 54 patients (N=6 with POD).** Effect sizes for the association of POD with postoperative relative SWI intensity (dependent variable) are given. See table S6 for a description of the sample. Abbreviations: B, regression coefficient; CI, confidence interval

| | **Reduced model[a]** | | | **Extended model[b]** | | |
|---|---|---|---|---|---|---|
| **ROI** | *B* | *90% CI* | *Partial $R^2$* | *B* | *90% CI* | *Partial $R^2$* |
| **Basal ganglia** | -0.029 | -0.065-0.006 | 0.037 | -0.027 | -0.060-0.006 | 0.038 |
| **Hippocampus** | 0.001 | -0.022-0.025 | <0.001 | 0.002 | -0.019-0.023 | <0.001 |
| **pBFCS** | -0.032 | -0.066-0.001 | 0.049 | -0.032 | -0.065-0.002 | 0.050 |

[a] Linear regression adjusted for duration of surgery (*df*=2/51)

[b] Linear regression adjusted for duration of surgery, age, sex, preoperative MMSE and ROI volume (*df*=6/47)

**Table S5. Demographic and clinical characteristics of the study sample with postoperative SWI data (*N*=34)**

| | | *N* | % |
|---|---|---|---|
| **Women** | | 17 | 50 |
| **ISCED** | **Level 1-2** | 2 | 6 |
| | **Level 3-4** | 14 | 45 |
| | **Level 5-6** | 15 | 48 |
| **Intrathoracic, -abdominal or -pelvic surgery** | | 5 | 15 |
| **ASA Physical Status** | **I** | 1 | 3 |
| | **II** | 22 | 65 |
| | **III** | 11 | 32 |
| **(Pre-)Frailty** | | 13 | 39 |
| **Hazardous alcohol consumption (AUDIT)** | | 2 | 6 |
| **Anesthesia** | **Regional** | 1 | 3 |
| | **General + regional** | 11 | 32 |
| **ICU admission** | | 1 | 3 |
| **Postoperative pain**[a] | | 12 | 38 |
| **Postoperative complication** | | 14 | 38 |
| **POD** | | 3 | 9 |
| | **Median** | **Interquartile range** | **Min.-max. range** |
| **Age (y)** | 72 | 67-75 | 65-86 |
| **CCI (p)** | 1 | 0-2 | 2-5 |
| **Baseline MMSE (p)** | 29 | 28-30 | 25-30 |
| **Surgery duration (min)** | 138 | 106-180 | 56-404 |
| **Days in hospital** | 7 | 4-8 | 2-12 |

**[a] Positive screening from a compound assessment including the Non-visual Rating Scale (NRS), Behavioral Pain Scale (BPS and BPS-NI) and Critical Pain Observation Tool (CPOT) during the first seven postoperative days**

Abbreviations: ASA, American Society of Anesthesiologists; AUDIT, Alcohol Use Disorder Identification Test; CCI, Charlson Comorbidity Index; d, days; ISCED, International Standard Classification of Education; Max., Maximum; min, minutes; Min., Minimum; MMSE, Mini-mental Status Examination; p, points; y, years;

**Table S6. Demographic and clinical characteristics of the study sample with postoperative SWI data (*N*=54).**

| | | ***N*** | **%** |
|---|---|---|---|
| **Women** | | 24 | 44 |
| **ISCED** | **Level 1-2** | 5 | 12 |
| | **Level 3-4** | 19 | 44 |
| | **Level 5-6** | 19 | 44 |
| **Intrathoracic, -abdominal or -pelvic surgery** | | 20 | 37 |
| **ASA Physical Status** | **I** | 2 | 4 |
| | **II** | 36 | 67 |
| | **III** | 16 | 30 |
| **(Pre-)Frailty** | | 16 | 31 |
| **Hazardous alcohol consumption (AUDIT)** | | 3 | 6 |
| **Anesthesia** | **Regional** | 1 | 2 |
| | **General + regional** | 17 | 31 |
| **ICU admission** | | 6 | 11 |
| **Postoperative pain**[a] | | 19 | 37 |
| **Postoperative complication** | | 24 | 44 |
| **POD** | | 6 | 11 |
| | **Median** | **Interquartile range** | **Min.-max. range** |
| **Age (y)** | 72 | 68-75 | 65-86 |
| **CCI (p)** | 2 | 0-2 | 0-7 |
| **Baseline MMSE (p)** | 29 | 28-30 | 25-30 |
| **Surgery duration (min)** | 160 | 107-246 | 56-425 |
| **Days in hospital** | 7 | 4-8 | 2-23 |
| [a] **and abbreviations: see table S5** | | | |

Acosta-Cabronero, J., Betts, M. J., Cardenas-Blanco, A., Yang, S., & Nestor, P. J. (2016). In Vivo MRI Mapping of Brain Iron Deposition across the Adult Lifespan. *J Neurosci*, *36*(2), 364-374. https://doi.org/10.1523/JNEUROSCI.1907-15.2016

Betts, M. J., Acosta-Cabronero, J., Cardenas-Blanco, A., Nestor, P. J., & Duzel, E. (2016). High-resolution characterisation of the aging brain using simultaneous quantitative susceptibility mapping (QSM) and R2* measurements at 7T. *Neuroimage*, *138*, 43-63. https://doi.org/10.1016/j.neuroimage.2016.05.024

Burgetova, R., Dusek, P., Burgetova, A., Pudlac, A., Vaneckova, M., Horakova, D., Krasensky, J., Varga, Z., & Lambert, L. (2021). Age-related magnetic susceptibility changes in deep grey matter and cerebral cortex of normal young and middle-aged adults depicted by whole brain analysis. *Quant Imaging Med Surg*, *11*(9), 3906-3919. https://doi.org/10.21037/qims-21-87

Haacke, E. M., Cheng, N. Y., House, M. J., Liu, Q., Neelavalli, J., Ogg, R. J., Khan, A., Ayaz, M., Kirsch, W., & Obenaus, A. (2005). Imaging iron stores in the brain using magnetic resonance imaging. *Magn Reson Imaging*, *23*(1), 1-25. https://doi.org/10.1016/j.mri.2004.10.001
